# Respiratory variation in mitral and tricuspid inflow velocity using real-time phase-contrast cardiovascular magnetic resonance - normal values and reproducibility

**DOI:** 10.1101/2023.07.31.23293410

**Authors:** Simon Thalén, Björn Wieslander, Einar Heiberg, Peder Sörensson, Daniel Giese, Andreas Sigfridsson, Martin Ugander

## Abstract

Ventricular interdependence occurs when pressure changes in one cardiac chamber affect the other, which can be exaggerated in cardiac pathology. The aim of this study was to develop a more automated, clinically accessible and freely available method to measure ventricular interdependence as respiratory variation in mitral and tricuspid peak early velocities. Patients without pericardial effusion or thickening (n=26, median age 55 years, 31% female), one patient with constrictive pericarditis, one patient with 35 mm pericardial effusion and healthy volunteers (n=13, median age 52 years, 76% female) underwent real-time phase-contrast cardiovascular magnetic resonance (CMR) at 3T in a basal short-axis view during free breathing. Healthy volunteers were imaged twice. Respiratory variation in mitral and tricuspid peak early inflow velocities was calculated as (V_max_-V_min_)/V_max_. In patients without pathology and healthy volunteers, which were pooled (n=39), respiratory variation was 23±6% (upper normal limit 34%) for mitral velocities and 40±10% (upper normal limit 60%) for tricuspid velocities. Patients with pathology exhibited increased respiratory variation. Test-retest variability and inter-observer agreements were excellent. The proposed method for quantifying respiratory variation of mitral and tricuspid inflow velocities by CMR is feasible, reproducible and normal values are presented.

## Introduction

Ventricular interdependence, also known as interaction or coupling, occurs when an increased pressure difference between the atrium and ventricle on one side of the heart leads to a compensatory decrease in the pressure difference on the opposite side (1). Despite its clinical relevance, current methods for quantifying this phenomenon, particularly using real-time phase-contrast cardiovascular magnetic resonance (RT-PC CMR), are limited by manual and cumbersome post-processing. Ventricular interdependence is assessed in clinical practice using Doppler echocardiography or invasive cardiac catheterization, where the respiratory variation in velocity or pressure is measured (2).

An increase or exaggeration of ventricular interdependence can occur in several clinical settings, most notably pericardial effusion/cardiac tamponade (3) and constrictive pericarditis (4). During inspiration, negative intrapleural pressures and increased abdominal pressures increase the venous return and rate of filling of the right heart, with a subsequent decrease in the rate of filling of the left heart due to interventricular interdependence. During expiration, the opposite occurs. In the case of a pericardial effusion compressing the heart, the combined volume of all heart chambers is more limited, and the venous return to the right heart occurs at a greater expense of the left, illustrating a more prominent ventricular interdependence.

The pericardium can gradually stretch to harbor liters of pericardial effusion in the setting of slowly evolving disease processes, but a rapid increase can trigger cardiac tamponade within minutes in acute disease (5,6). Thus, assessment of effusion volume alone is inadequate to identify the impending onset of tamponade. Hemodynamic events, including right atrial collapse during ventricular systole, right ventricular early diastolic collapse, and increased ventricular interdependence, typically precede tamponade (5,7). Pulsus paradoxus is considered an important clinical sign of imminent tamponade and is defined as a decrease of over 10 mmHg in systolic blood pressure during inspiration, and is a consequence of ventricular interdependence (8–10). Respiratory changes in transvalvular peak early inflow velocities by Doppler echocardiography have been described as a surrogate marker for pulsus paradoxus (11) and flow velocity paradoxus (12). Furthermore, the diagnosis of constrictive pericarditis remains a challenge, and respiratory variation in mitral and tricuspid inflow velocities is considered a valuable tool for separating constrictive and restrictive physiologies (4,13–15).

Previous studies of ventricular interdependence have evaluated patients with constrictive pericarditis by quantifying the respiratory difference in maximal septal excursion normalized to biventricular diameter (16) or used real-time free-breathing CMR to calculate an index of ventricular interdependence (17). Moreover, RT-PC CMR has been employed to measure respiratory variation in transvalvular inflow velocities, showing excellent accuracy in identifying constrictive physiology (18). However, the quantification of respiratory variation or normalized differences in maximal septal excursion using RT-PC CMR has seen limited clinical adoption, partly due to the cumbersome and manual post-processing required.

The present study aimed to develop a more automated, clinically accessible and freely available method to measure ventricular interdependence by simultaneously quantifying the respiratory variation in mitral and tricuspid peak early inflow velocities using semi-automatic analysis of RT-PC CMR images, to derive normal reference values and determine test-retest variability.

## Materials and methods

### Study design

The present study is a prospective imaging study of patients who underwent CMR on clinical indications and volunteers who underwent CMR for research purposes. The primary outcomes were respiratory variation in mitral and tricuspid peak early inflow velocity and test-retest variability.

### Patients

CMR imaging occurred between June 2020 and December 2021 in the Department of Clinical Physiology, Karolinska Universitetsjukhuset, Stockholm, Sweden. A random selection was made of patients (n=27) who did not exhibit pericardial effusion, pericardial thickening, or septal bounce. Feasibility was illustrated by imaging one patient with constrictive pericarditis and one patient with a hemodynamically significant pericardial effusion measuring 35 mm, characterized by an above 25% respiratory variation in mitral inflow velocities by echocardiography. Healthy volunteers (n=15) were recruited by word of mouth. After each scanning session, volunteers exited the scanner, walked around it once, and were re-imaged using the same methodology.

### Image acquisition

CMR studies were conducted using a research sequence at 3T (MAGNETOM Skyra, Siemens Healthineers, Forchheim, Germany) in a basal short-axis view over a 30-second acquisition during free breathing. The typical image acquisition parameters included a repetition time of 3.7 ms, water excitation pulse with a flip angle of 10°, slice thickness of 8 mm, field of view of 360×266 mm², matrix of 208×135, velocity encoding of 150 cm/s, and shared velocity encoding enabled (19). Compressed sensing with an acceleration factor of 7.7 was used to achieve a temporal resolution of 47.8 ms (19).

RT-PC velocity-encoded imaging data were acquired with through-plane velocity encoding, with the slice positioned slightly apical to the atrioventricular plane at the tips of the valve leaflets in end diastole. Planning was performed on a four-chamber long-axis view (Figure 4C). A 30-second RT-PC dataset spanning multiple cardiac and respiratory cycles was obtained for each patient and volunteer. All subjects were in sinus rhythm during the CMR imaging.

### Image analysis

Image analysis was performed offline using an in-house-developed software plugin integrated with the Segment software program (Medviso AB, Lund, Sweden) (21). The plugin was developed using source code available for research within the MATLAB software environment (version 2019a, The MathWorks, Inc., Natick, Massachusetts, United States). Phase offset errors were corrected using stationary tissue background correction and established methods (20).

The plugin lets users manually define regions of interest (ROIs) encompassing the mitral and tricuspid orifices that are subsequently propagated across all time frames. The ROIs can be reviewed frame by frame to ensure accurate placement. The negative flow velocity component within the mitral ROI during systole, representing part of the aortic outflow, is utilized to automatically segment the data into individual cardiac cycles by identifying the peak negative flow, part of the aortic outflow, as the divider between cycles. The visibility of the dividers can be toggled, and each divider can be adjusted manually if needed.

Spectral plots of mitral and tricuspid velocities are generated, and the mean velocity of pixels exceeding the 95th percentile is used to derive a velocity-time curve for identifying peak early inflow velocities. The peak early inflow velocity is automatically identified as the first local maxima of a given cardiac cycle with a value exceeding the mean plus one standard deviation of the positive velocity in each timeframe. Individual peak velocities can be manually adjusted or deleted. A time-motion graph, with time on the horizontal axis and voxels from a vertical line that can be selected from the magnitude image on the vertical axis, allows for visual tracking of the lung-diaphragm interface. Figure 2 shows an example of the interface of the plugin and the result from a patient with normal physiology.

**Figure 1.**
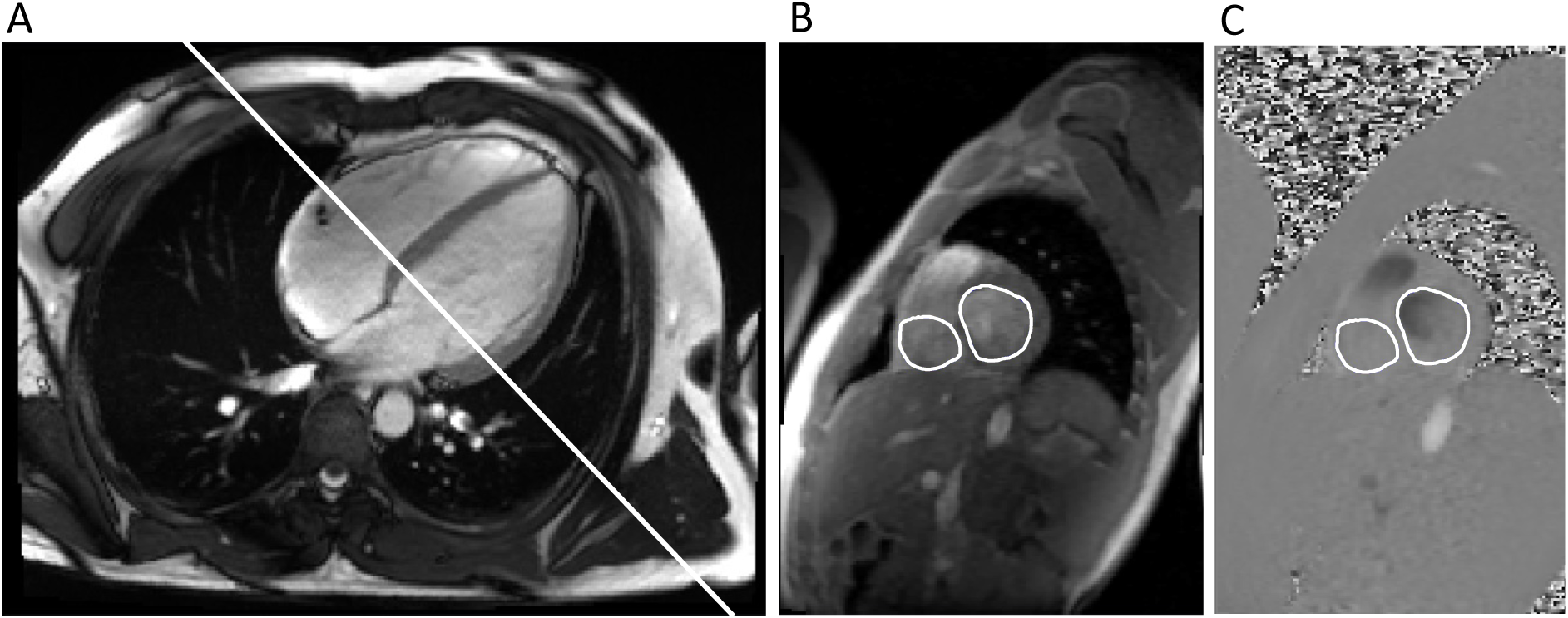
A four-chamber long-axis cine acquisition in end-diastole showing the slice orientation of the through-plane RT-PC imaging (A). Regions of interest for the mitral and tricuspid valves are shown in magnitude and phase images (B and C).

**Figure 2.**
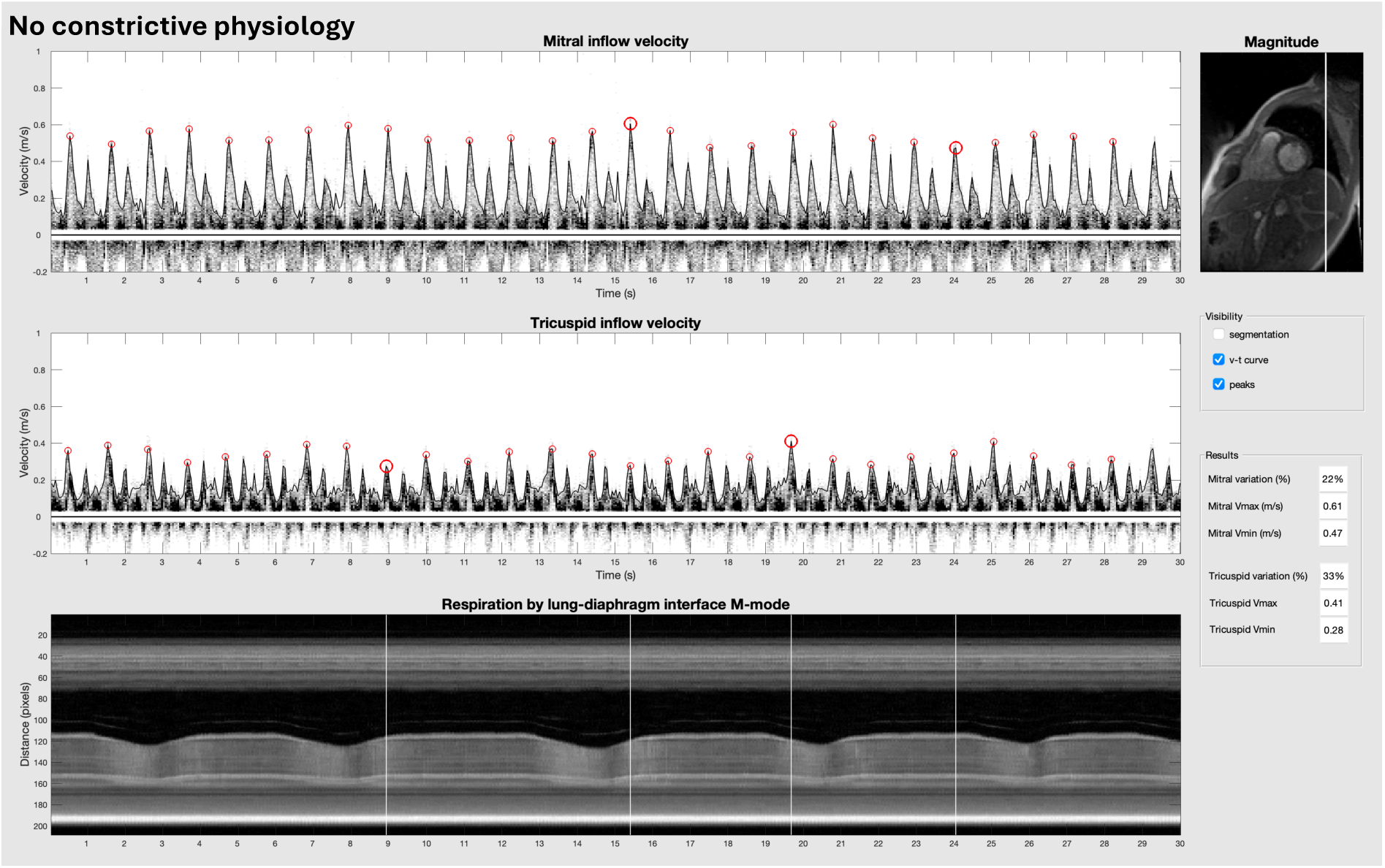
An example of a patient without constrictive physiology is shown. The two upper-left panels display spectral plots of all velocities, with each peak early inflow velocity marked by a red circle. The lower-left panel presents a time-motion graph, with time on the horizontal axis and voxels from a vertical line selected from the magnitude image (top right) on the vertical axis. This graph is analogous to M-mode echocardiography, allowing for visual tracking of the lung-diaphragm interface. In this graph, white lines indicate each mitral and tricuspid V_max_ and V_min_, enabling the user to confirm that each reported peak occurred during inspiration or expiration. The visibility of cardiac cycle segmentation, velocity-time curve, and peaks can be toggled in the middle-right panel and results presented in the bottom-right panel.

The respiratory variation in peak early inflow velocities was defined as the absolute change value as a percentage of the maximum peak early inflow velocity as described below in Eq. 1.

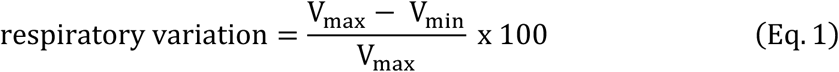

Alternative definitions used in prior literature are summarized in Appendix A for comparison.

### Statistical analysis

The normality of distributions was assessed both visually using Q-Q plots and the Shapiro-Wilk test. Data are reported as median [interquartile range] or mean ± SD as appropriate. Normal reference ranges were calculated as the mean ± 1.96 times the standard deviation, giving the 95% upper and lower normal limits. Intraclass correlation (ICC) estimates were calculated using a single rating (k=1), absolute agreement, and a 2-way random-effects model (22). All statistical calculations were performed using the software package R (R Core Team 2020, Vienna, Austria).

## Results

One patient was excluded because of inaccurate slice planning and two volunteer measurements were excluded due to poor image quality in at least one of the two acquisitions, leaving a total of 26 patients and 13 healthy volunteers included in the final analysis. Patient and volunteer characteristics are summarized in Table 1.

**Table 1.** Clinical characteristics. Clinical characteristics and CMR findings in 26 patients without increased pericardial thickening or effusion and 13 healthy volunteers.

| Characteristic |  |
| --- | --- |
| Mitral respiratory variation*, % | 23 ± 6 |
| Tricuspid respiratory variation*, % | 41 ± 10 |
| Maximum mitral early inflow velocity, m/s | 0.57 ± 0.17 |
| Minimum mitral early inflow velocity, m/s | 0.44 ± 0.15 |
| Maximum tricuspid early inflow velocity, m/s | 0.40 ± 0.15 |
| Minimum tricuspid early inflow velocity, m/s | 0.24 ± 0.11 |
| Left ventricular ejection fraction†, % | 56 (47-58) |
| Age, years | 50 (36-63) |
| Female, % | 46 |

Continuous data is given as mean ± SD, median (interquartile range) or number (%).
\* Calculated as described in Eq. 1. † The ejection fraction of the healthy volunteers was assessed visually and found to be normal.

As the respiratory variation in mitral and tricuspid peak early inflow velocities did not differ between patients and healthy volunteers analyzed by operator 1 (p=0.90 and p=0.27, respectively), the data were pooled to enhance statistical robustness.

The pooled respiratory variation in mitral and tricuspid peak early inflow velocity in patients without constrictive pericarditis or pericardial effusion and healthy volunteers was 23±6% (upper normal limit 34%) and 40±10% (upper normal limit 60%), respectively.

All mitral peak early maximum velocities occurred during expiration and the minimum velocities during inspiration. Conversely, all tricuspid peak early maximum velocities occurred during inspiration and minimum velocities during expiration.

Two acquisitions from each of the remaining 13 healthy volunteers were analyzed in random order by two operators (ST and BW) blind to the subject identities.

The mean difference±standard deviation in test-retest differences for respiratory variation was −1±5% for mitral and −1±5% for tricuspid peak inflow velocities. Test-retest repeatability was assessed using operator 1 results, and the test-retest ICC was 0.73 for mitral and 0.89 for tricuspid respiratory variation. The median [interquartile range] in inter-observer differences for respiratory variation was 0 [0–0] % for mitral and 0 [-0.7–0.2] % for tricuspid peak inflow velocities.

The inter-observer results, which were pooled (n=26), showed an inter-observer ICC of 0.98 for mitral and 0.99 for tricuspid respiratory variation. Bland-Altman plots of the test-retest and reproducibility results are presented in Figure 5.

**Figure 3.**
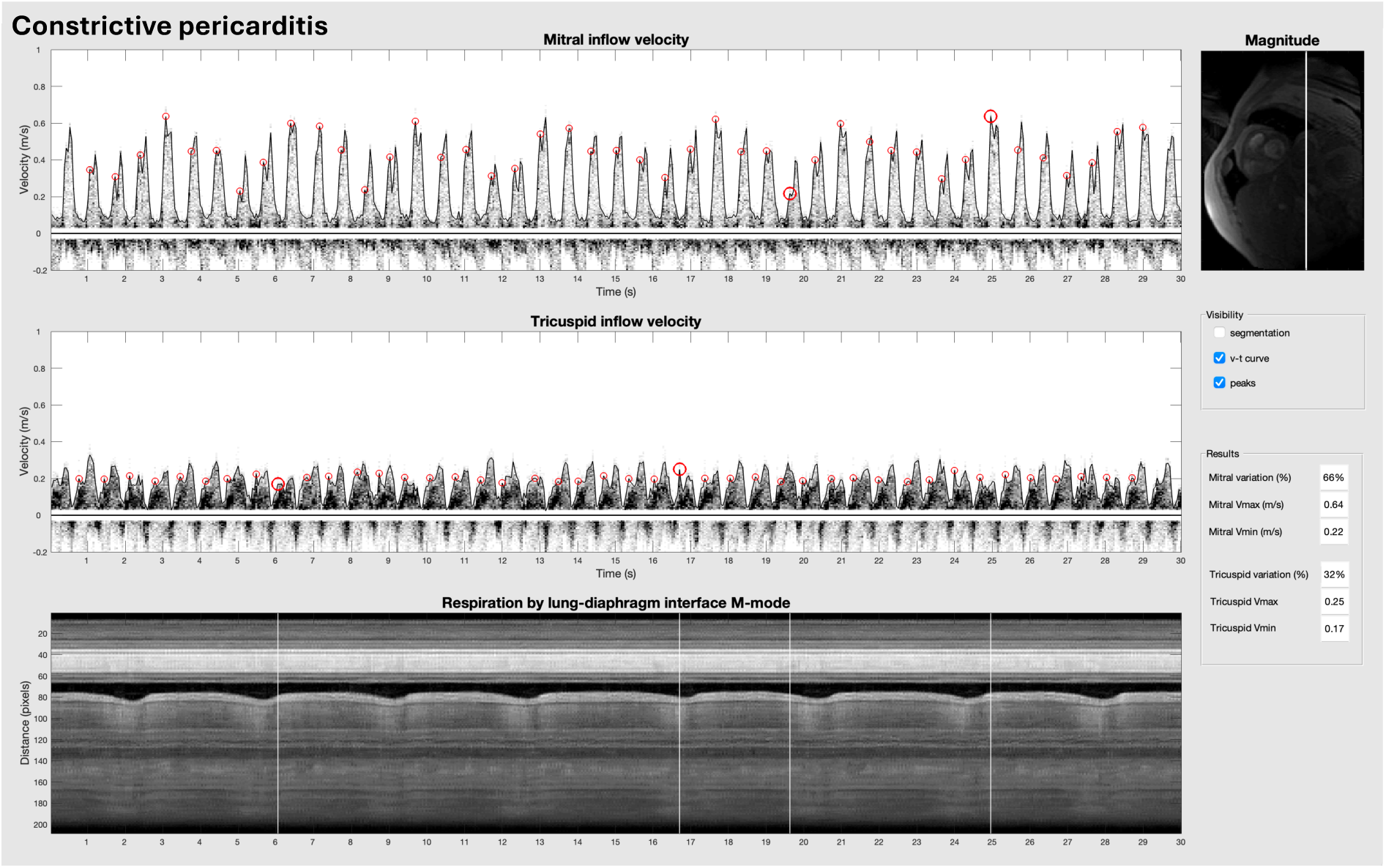
An example of a patient with constrictive pericarditis with mitral and tricuspid variation in peak early inflow velocities of 66% and 32%.

**Figure 4.**
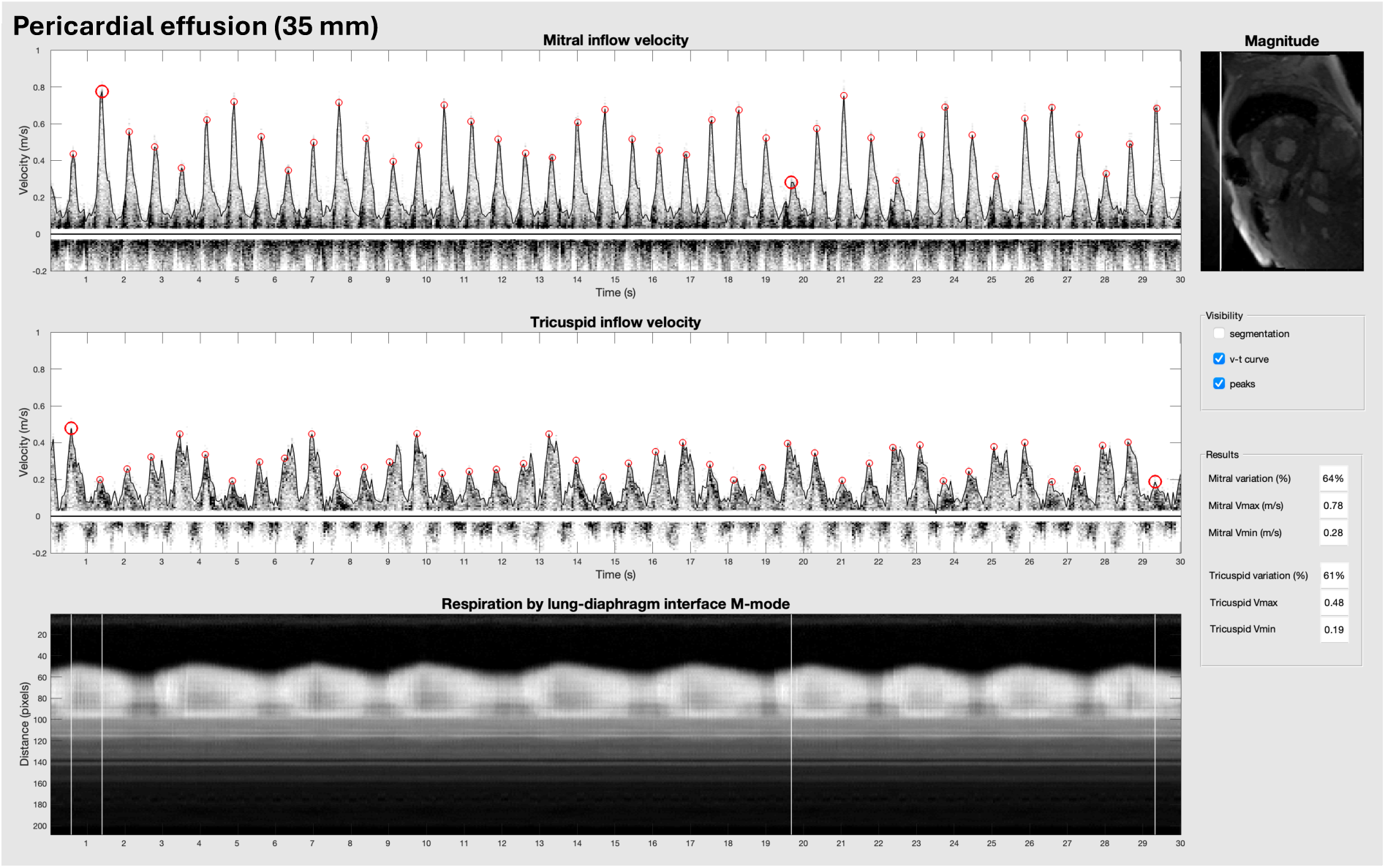
An example of a patient with 35 mm pericardial effusion with mitral and tricuspid variation in peak early inflow velocities of 66% and 61%.

**Figure 5.**
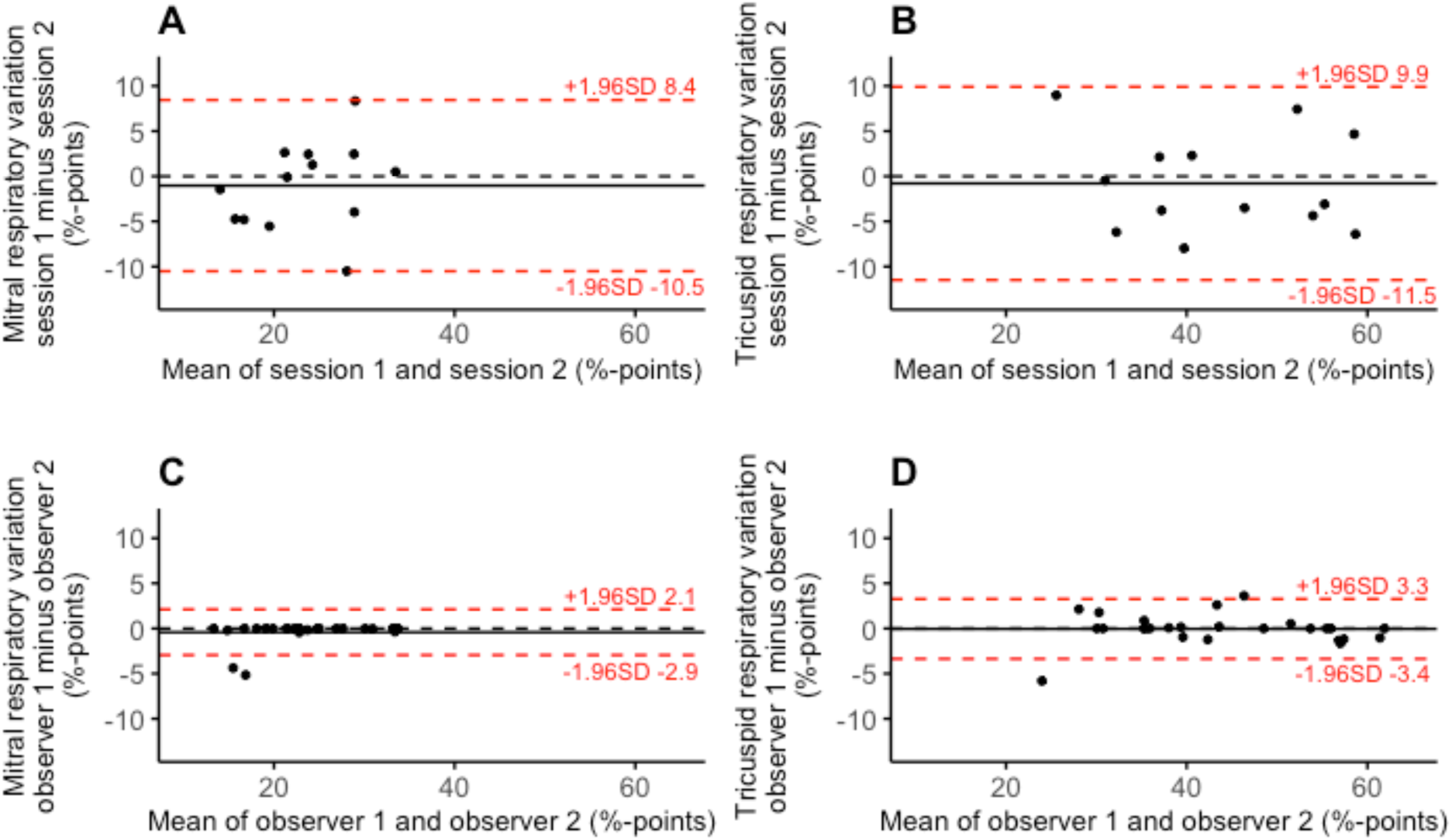
Bland-Altman plots of bias and limits of agreement (in red) of the difference between test-retest results for mitral velocity (top-left), test-retest for tricuspid velocity (top-right), inter-observer results for mitral velocity (bottom-left), and inter-observer results for tricuspid velocity (bottom-right).

The patient with constrictive pericarditis, presented in Figure 3, had a respiratory variation in mitral and tricuspid peak early inflow of 66% and 32%.

The patient with a 35 mm pericardial effusion, presented in Figure 4, had a respiratory variation in mitral and tricuspid peak early inflow velocities of 64% and 61%.

## Discussion

The present study shows clinical feasibility and presents normal values for respiratory variation in mitral and tricuspid peak early inflow velocities measured using RT-PC CMR. The normal values of the current study are higher than those obtained with Doppler echocardiography. In the first Doppler study on respiratory variation in cardiac tamponade and pericardial effusion, almost no respiratory variation in mitral inflow velocity in normal subjects could be detected (3). Likewise, a later study reported nearly no respiratory variation in velocity-time integrals of mitral inflow in normal subjects (25).

Higher values were observed when comparing results using the same definition of maximal respiratory variation as a prior study using RT-PC CMR (as defined by Eqs 3 and 4 in Appendix A). Several factors are likely contributing to this. In the previous study, peak early inflow velocities were considered solely from the first heartbeat of each respiratory phase. In contrast, the current approach considers all peak early inflow velocities across all available respiratory cycles, utilizing the maximum and minimum velocities observed throughout the full range of respiration. The longer acquisition time in the present study, 30 seconds, compared to between 10 and 20 seconds in the previous study, increases the likelihood of capturing mitral or tricuspid peak early velocities that align with end inspiration or end expiration, thereby amplifying the observed variation.

Additionally, the higher temporal resolution of the sequence used in the current study may have resulted in a velocity-time curve that more accurately captured high and low peak velocities, leading to a further increase. Separating peak early from peak atrial inflow for each cardiac cycle can be challenging, but less so using a high temporal resolution. Measuring the peak atrial, instead of peak early inflow velocities, would likely result in lower respiratory variation as atrial contraction is less affected by respiration. The risk is greater for the tricuspid valve, whose inflow velocity is lower and, therefore, more challenging to characterize. Simultaneous mitral and tricuspid flow velocity measurements are possible using RT-PC. Notably, the results of the present study demonstrate a relatively high normal upper limit of tricuspid respiratory variation, which calls into question the value of measuring tricuspid respiratory variation in evaluating ventricular interdependence using this method.

Given the variability in peak early inflow velocities among patients, respiratory variation is typically expressed as a percentage, calculated from the positive, negative, or absolute difference between inspiratory (V_insp_) and expiratory (V_exp_) velocities, with either V_insp_ or V_exp_ as the denominator. For mitral velocities, using V_insp_ as the denominator reflects an expiratory increase relative to V_insp_, while using V_exp_ reflects an inspiratory decrease relative to V_exp_. The reverse applies to tricuspid velocities, where V_insp_ corresponds to an expiratory decrease and V_exp_ to an inspiratory increase. The choice of denominator is often implied rather than explicitly stated in the literature. Most authors conceptualize mitral respiratory variation as an inspiratory decrease and tricuspid variation as an inspiratory increase (2,3,26–31), aligning with the definition of pulsus paradoxus as a systolic pressure drop during inspiration (8) and the current guidelines from the American Society for Echocardiography (2). This introduces an asymmetry between the mitral and tricuspid variation because change expressed as a percent increase will always be greater than the same change expressed as a percentage decrease by simple arithmetic.

Some studies report maximal respiratory change as the expiratory increase in mitral inflow and expiratory decrease in tricuspid inflow (18), while others define respiratory variation relative to apnea (4,32) or as an absolute change in velocities (33). As highlighted earlier (34) and supported by our study, results vary markedly depending on the denominator used, particularly for tricuspid velocities. For simplicity and consistency, we adopted Eq. 1 as the definition, which is straightforward, symmetrical, and aligned with our automated analysis approach. This approach is valid if all mitral peak early maximum velocities occur during expiration and the minimum velocities during inspiration, and all tricuspid peak early maximum velocities occur during inspiration and the minimum velocities during expiration. This was verified using visual tracking of the lung-diaphragm interface in a time-motion graph as part of the software tool. Full results using alternative published definitions are available in Appendix A.

The present study also evaluated the test-retest repeatability and interobserver reproducibility of the method. To date, no studies have investigated test-retest repeatability or interobserver reproducibility of ventricular interdependence measurements using invasive pressure assessments, respiratory variation in mitral and tricuspid early filling velocities via pulsed-wave Doppler echocardiography, or phase-contrast CMR, precluding any direct comparison of these findings.

The low variability between observers in our study is likely due to the inherent robustness of automated peak early inflow velocity measurements against variations in the geometry of the ROIs. Given that peak velocities consistently fall within the ROI and that any potential inclusion of noise from regions outside the myocardium would have been readily apparent, the time-velocity curves generated by the method are unaffected by minor differences in ROI placement. This finding indicates that differences between exams have a greater impact on measurement results than differences between observers, consistent with previous research on test-retest and inter-observer variability in diastolic function measurements in echocardiography (35).

The method presented in the present study differs from a purely morphological imaging method such as 2D echocardiography or cine CMR, which are used to quantify the size of the effusion or thickness of the pericardium in millimeters. The method introduced in this study assesses a physiological outcome resulting from changes in the cardiac pressure-volume relationship during respiration. It has been found that incidental findings of clinically meaningful pericardial effusion occurred in 0.8% of CMR exams (36). The capability to instantly assess ventricular interdependence to determine its hemodynamic significance would be beneficial. In the case of constrictive pericarditis, an evaluation of ventricular interdependence is also valuable, as 18% of surgically proven cases have been found to lack pericardial thickening and thus require hemodynamic evaluation (37).

## Strengths and limitations

The freely available and semi-automatic methodology for analyzing RT-PC images is a strength of the present study. Besides drawing two ROIs, evaluation using the software plugin typically requires no further user input. The velocity-time curve is unambiguously calculated, reducing the risk of confirmation bias.

Historically, measurements of respiratory variation in mitral and tricuspid velocities have relied upon observations of chest movements (25), thermistor recordings (3,38), or transducer recordings (24) to track respiratory motion, and manual calculations on paper tracings to estimate the velocity-time curve. In contrast, the present study used the lung– diaphragm interface, clearly identifiable in the magnitude component of short-axis images, to visualize and demarcate inspiration and expiration without requiring extra equipment.

The study also has limitations. First, only a single patient with constrictive pericarditis and one with a hemodynamically significant pericardial effusion were included, precluding any inference of diagnostic accuracy; the findings should be interpreted primarily as a feasibility and normal-values study. Second, the cohort comprised middle-aged patients already referred for CMR and healthy volunteers of similar age, limiting generalizability to younger individuals, pediatric patients, or those with arrhythmias, lung disease, or altered loading conditions. Third, all examinations were performed at a single center using a single vendor platform, and interobserver reproducibility was assessed only between experienced operators; reproducibility across vendors and among less experienced readers remains to be determined. A direct comparison to Doppler echocardiography would have been of interest but was outside the scope of the study. Finally, respiratory loading conditions and through-plane valve motion may confound measurements. Cardiac motion during respiration causes displacement of the imaging plane, and the mitral and tricuspid valves are particularly susceptible to such motion artifacts (39,40). This problem is not limited to RT-PC CMR imaging. Doppler echocardiography is commonly used for studies of respiratory variation in transvalvular inflow velocities and is also vulnerable to displacement during breathing. The effect of in-plane displacement due to respiration was minimized by ensuring that the mitral and tricuspid orifices were fully contained within the regions of interest (ROIs) across all timeframes. Future studies should prospectively evaluate diagnostic performance compared to Doppler echocardiography and, ideally, also cardiac catheterization, in larger and more diverse patient populations, assess reproducibility across centers and vendors, and explore applications beyond pericardial disease, such as pulmonary hypertension, right ventricular overload, or diastolic dysfunction. The potential for inline automated analysis during CMR acquisition and in-plane measurements of respiratory variation in peak early inflow velocities, including a simultaneous estimation of septal displacement, warrants further investigation.

## Conclusions

The present study reports normal values and demonstrates that semi-automatic analysis of RT-PC CMR flow measurement is feasible and can be used to evaluate ventricular interdependence by measuring respiratory variation in mitral and tricuspid early inflow velocities. The proposed method has an acceptable test–retest repeatability, and the interobserver reproducibility is excellent. While the results indicate preliminary clinical feasibility, the limited number of patients with constrictive pericarditis or significant pericardial effusion precludes conclusions regarding diagnostic accuracy. Further evaluation of its utility for diagnosing and managing conditions like constrictive pericarditis and pericardial effusion is justified. The method is made freely available as open-source software.

## Data Availability

All data produced in the present study are available upon reasonable request to the authors

https://github.com/simonthalen/segment-open-and-respiratory-variation

## Declarations

### Ethics approval and consent to participate

The patient component of the present study was a retrospective study of clinically acquired imaging data. All patients provided written informed consent prior to enrolment. The patients were already scheduled to undergo CMR imaging as part of their clinical examination. All patient data were anonymized prior to analysis to ensure the integrity of the patient’s identities. Healthy subjects were prospectively recruited following written informed consent. Approval from the Stockholm Regional Ethics Board was obtained (Number 2011/1077-31/3).

### Consent for publication

Written informed consent was obtained from patients for publication of their individual details on a group level and anonymized images in this manuscript. The consent form is held in the patient’s clinical notes and is available for review by the Editor-in-Chief.

### Software and data availability

The plugin used is part of the software Segment, which is freely available for research purposes from https://medviso.com/. Platform-independent MATLAB source code, R-notebook and data used for statistical calculations and generation of Figure 5 are available as at https://github.com/simonthalen/segment-open-and-respiratory-variation with release DOI: doi.org/10.5281/zenodo.17179338. All data produced in the present study are available from the corresponding author upon reasonable request.

### Author contributions

Simon Thalén contributed to the design, data acquisition and analysis, and manuscript drafting. Björn Wieslander and Einar Heiberg contributed to the data analysis of the work.

Andreas Sigfridsson, Daniel Giese, Henrik Engblom and Peder Sörensson contributed to the data acquisition. Martin Ugander contributed to the design of the study, data interpretation, and drafting the manuscript. All authors gave final approval of the version to be published and agreed to be accountable for all aspects of the work in ensuring that questions related to the accuracy or integrity of any part of the work are appropriately investigated and resolved.

## Acknowledgments

We acknowledge the support provided by the CMR technologists at the Department of Clinical Physiology, Karolinska University Hospital.

## Appendix A

Several different equations have been proposed in the literature to quantify respiratory variation in transvalvular peak early inflow velocities. The respiratory variation in peak early inflow velocities is defined as the absolute change value as a percentage of the maximum peak early inflow velocity, as described in Eq. 1.

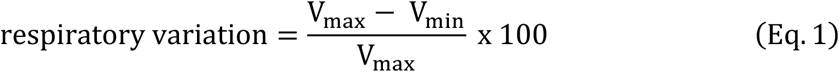

In this study, we used Eq. 1, but results using other definitions are also provided for comparison. A second definition as previously described (2) was used as follows:

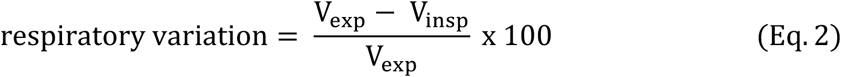

Finally, a third definition as previously described (18) was used as follows:

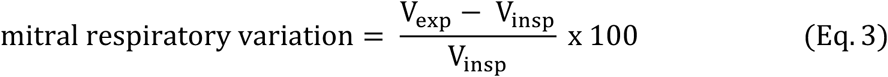

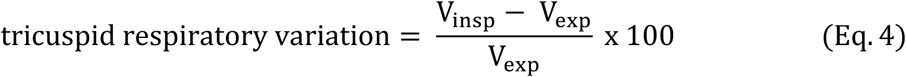

The pooled respiratory variation in mitral and tricuspid peak early inflow velocity using Eq. 1 in patients without constrictive pericarditis or pericardial effusion and healthy volunteers was 23±6% (upper normal limit 34%) and 40±10% (upper normal limit 60%), respectively. Using the alternative definitions such as Eq. 2, the respiratory variation in mitral peak early inflow velocities was, for arithmetical reasons identical, and the tricuspid peak early inflow velocity was −74±33% (lower normal limit −139%). Using Eqs. 3 and 4, the respiratory variation in mitral peak early inflow velocities was 30±10% (upper normal limit 50%) and tricuspid peak early inflow velocities 74±33% (upper limit 139%).

The patient with constrictive pericarditis, presented in Figure 3, had a respiratory variation in mitral and tricuspid peak early inflow velocities using the definitions in Eq. 1 of 66% and 32%, Eq. 2 of 66% and −47 %, and Eqs. 3 and 4, 191% and 47%, respectively. The patient with a 35 mm pericardial effusion, presented in Figure 4, had a respiratory variation in mitral and tricuspid peak early inflow velocities using the definitions in Eq. 1 of 64% and 61%, Eq. 2 of 64% and −155%, and Eqs. 2 and 3, 177% and 155%, respectively.

## References

1. Bove AA, Santamore WP. Ventricular interdependence. Prog Cardiovasc Dis. 1981;23(5):365–388. doi:10.1016/0033-0620(81)90022-0

2. Klein AL, Abbara S, Agler DA, et al. American Society of Echocardiography Clinical Recommendations for Multimodality Cardiovascular Imaging of Patients with Pericardial Disease. J Am Soc Echocardiogr. 2013;26(9):965–1012.e15. doi:10.1016/j.echo.2013.06.023

3. Appleton CP, Hatle LK, Popp RL. Cardiac tamponade and pericardial effusion: respiratory variation in transvalvular flow velocities studied by Doppler echocardiography. J Am Coll Cardiol. 1988;11(5):1020–1030.

4. Hatle LK, Appleton CP, Popp RL. Differentiation of constrictive pericarditis and restrictive cardiomyopathy by Doppler echocardiography. Circulation. 1989;79(2):357–370.

5. Shabetai R. Pericardial effusion: haemodynamic spectrum. Heart. 2004;90(3):255–256. doi:10.1136/hrt.2003.024810

6. Saito Y, Donohue A, Attai S, et al. The Syndrome of Cardiac Tamponade with “Small” Pericardial Effusion. Echocardiography. 2008;25(3):321–327. doi:10.1111/j.1540-8175.2007.00567.x

7. Roy CL, Minor MA, Brookhart MA, Choudhry NK. Does this patient with a pericardial effusion have cardiac tamponade? JAMA. 2007;297(16):1810–1818.

8. Dornhorst AC, Howard P, Leathart GL. Pulsus paradoxus. Lancet Lond Engl. 1952;1(6711):746–748.

9. Curtiss EI, Reddy PS, Uretsky BF, Cecchetti AA. Pulsus paradoxus: Definition and relation to the severity of cardiac tamponade. Am Heart J. 1988;115(2):391–398. doi:10.1016/0002-8703(88)90487-5

10. Hamzaoui O, Monnet X, Teboul JL. Pulsus paradoxus. Eur Respir J. 2013;42(6):1696–1705. doi:10.1183/09031936.00138912

11. Alerhand S, Carter JM. What echocardiographic findings suggest a pericardial effusion is causing tamponade? Am J Emerg Med. 2019;37(2):321–326. doi:10.1016/j.ajem.2018.11.004

12. Shyy W, Knight RS, Kornblith A, Teismann NA. Point-of-Care Diagnosis of Cardiac Tamponade Identified by the Flow Velocity Paradoxus. J Ultrasound Med. 2017;36(11):2197–2201. doi:10.1002/jum.14251

13. Nishimura RA. Constrictive pericarditis in the modern era: a diagnostic dilemma. Heart. 2001;86(6):619–623.

14. Oh JK, Hatle LK, Seward JB, et al. Diagnostic role of Doppler echocardiography in constrictive pericarditis. J Am Coll Cardiol. 1994;23(1):154–162.

15. Hancock EW. Differential diagnosis of restrictive cardiomyopathy and constrictive pericarditis. Heart. 2001;86(3):343–349. doi:10.1136/heart.86.3.343

16. Francone M, Dymarkowski S, Kalantzi M, Bogaert J. Real-time cine MRI of ventricular septal motion: A novel approach to assess ventricular coupling. J Magn Reson Imaging. 2005;21(3):305–309. doi:10.1002/jmri.20259

17. Anavekar NS, Wong BF, Foley TA, et al. Index of biventricular interdependence calculated using cardiac MRI: a proof of concept study in patients with and without constrictive pericarditis. Int J Cardiovasc Imaging. 2013;29(2):363–369. doi:10.1007/s10554-012-0101-x

18. Thavendiranathan P, Verhaert D, Walls MC, et al. Simultaneous Right and Left Heart Real-Time, Free-Breathing CMR Flow Quantification Identifies Constrictive Physiology. JACC Cardiovasc Imaging. 2012;5(1):15–24. doi:10.1016/j.jcmg.2011.07.010

19. Lustig M, Donoho D, Pauly JM. Sparse MRI: The application of compressed sensing for rapid MR imaging. Magn Reson Med. 2007;58(6):1182–1195. doi:10.1002/mrm.21391

20. Nickander J, Lundin M, Abdula G, et al. Stationary tissue background correction increases the precision of clinical evaluation of intra-cardiac shunts by cardiovascular magnetic resonance. Sci Rep. 2020;10(1):5053. doi:10.1038/s41598-020-61812-7

21. Heiberg E, Sjögren J, Ugander M, Carlsson M, Engblom H, Arheden H. Design and validation of Segment--freely available software for cardiovascular image analysis. BMC Med Imaging. 2010;10:1. doi:10.1186/1471-2342-10-1

22. Koo TK, Li MY. A Guideline of Selecting and Reporting Intraclass Correlation Coefficients for Reliability Research. J Chiropr Med. 2016;15(2):155–163. doi:10.1016/j.jcm.2016.02.012

23. Bland JM, Altman D. Statistical methods for assessing agreement between two methods of clinical measurement. The lancet. 1986;327(8476):307-310.

24. Bland JM, Altman DG. Measuring agreement in method comparison studies. Stat Methods Med Res. 1999;8(2):135–160. doi:10.1177/096228029900800204

25. Leeman DE, Levine MJ, Come PC. Doppler echocardiography in cardiac tamponade: exaggerated respiratory variation in transvalvular blood flow velocity integrals. J Am Coll Cardiol. 1988;11(3):572–578.

26. Ginghina C, Beladan C, Iancu M, Calin A, Popescu BA. Respiratory Maneuvers in Echocardiography: A Review of Clinical Applications. Vol 7.; 2009. doi:10.1186/1476-7120-7-42

27. Burstow DJ, Oh JK, Bailey KR, Seward JB, Tajik AJ. Cardiac Tamponade: Characteristic Doppler Observations. Mayo Clin Proc. 1989;64(3):312–324. doi:10.1016/S0025-6196(12)65251-3

28. Simeonidou E, Hamouratidis N, Tzimas K, Tsounos J, Roussis S. Respiratory variation in mitral flow velocity in pericardial effusion and cardiac tamponade. Angiology. 1994;45(3):213–218.

29. Zhang S, Kerins DM, Iii BFB. Doppler Echocardiography in Cardiac Tamponade and Constrictive Pericarditis. Echocardiography. 1994;11(5):507–521. doi:10.1111/j.1540-8175.1994.tb01092.x

30. Kim JS, Ha JW, Im E, et al. Effects of pericardiectomy on early diastolic mitral annular velocity in patients with constrictive pericarditis. Int J Cardiol. 2009;133(1):18–22. doi:10.1016/j.ijcard.2007.11.064

31. Appleton C, Gillam L, Koulogiannis K. Cardiac Tamponade. Cardiol Clin. 2017;35(4):525–537. doi:10.1016/j.ccl.2017.07.006

32. Picard MH, Sanfilippo AJ, Newell JB, Rodriguez L, Guerrero JL, Weyman AE. Quantitative relation between increased intrapericardial pressure and Doppler flow velocities during experimental cardiac tamponade. J Am Coll Cardiol. 1991;18(1):234–242. doi:10.1016/s0735-1097(10)80245-1

33. Gonzalez MS, Basnight MA, Appleton CP. Experimental pericardial effusion: relation of abnormal respiratory variation in mitral flow velocity to hemodynamics and diastolic right heart collapse. J Am Coll Cardiol. 1991;17(1):239–248.

34. Siniorakis E, Arvanitakis S, Zarreas E, Barlagiannis D, Skandalakis N, Karidis C. Calculating the respiratory flow velocity fluctuations in pericardial diseases. Int J Cardiol. 2010;145(1):92–93. doi:10.1016/j.ijcard.2009.05.052

35. Palmieri V, Arezzi E, Sabatella M, Celentano A. Interstudy reproducibility of parameters of left ventricular diastolic function: a Doppler echocardiography study. J Am Soc Echocardiogr Off Publ Am Soc Echocardiogr. 2003;16(11):1128–1135. doi:10.1067/S0894-7317(03)00641-2

36. McKenna DA, Laxpati M, Colletti PM. The Prevalence of Incidental Findings at Cardiac MRI. Open Cardiovasc Med J. 2008;2:20–25. doi:10.2174/1874192400802010020

37. Talreja DR, Edwards WD, Danielson GK, et al. Constrictive Pericarditis in 26 Patients With Histologically Normal Pericardial Thickness. Circulation. 2003;108(15):1852–1857. doi:10.1161/01.CIR.0000087606.18453.FD

38. Wayne VS, Bishop RL, Spodick DH. Dynamic effects of pericardial effusion without tamponade. Respiratory responses in the absence of pulsus paradoxus. Br Heart J. 1984;51(2):202–204.

39. Seemann F, Heiberg E, Carlsson M, et al. Valvular imaging in the era of feature-tracking: A slice-following cardiac MR sequence to measure mitral flow. J Magn Reson Imaging. 2020;51(5):1412–1421. doi:10.1002/jmri.26971

40. Fidock B, Archer G, Barker N, et al. Standard and emerging CMR methods for mitral regurgitation quantification. Int J Cardiol. 2021;331:316–321. doi:10.1016/j.ijcard.2021.01.066

